# Association of Clonal Hematopoiesis with Total and Cause-Specific Mortality Among Older Women

**DOI:** 10.64898/2026.05.28.26354392

**Authors:** Annie Chang, Daniel Ezzat, Md Mesbah Uddin, Yash Pershad, Jason M. Collins, Jacob O. Kitzman, Siddhartha Jaiswal, Pinkal Desai, Aladdin H. Shadyab, Garnet L. Anderson, Ramon Casanova, Robert Wallace, Jean Wactawski-Wende, Alexander G. Bick, Pradeep Natarajan, Charles Kooperberg, Michael J. LaMonte, Eric A. Whitsel, JoAnn E. Manson, Alexander P. Reiner, Michael C. Honigberg

**Author notes:** Indicates shared authorship (Annie Chang, Daniel Ezzat). **Address for correspondence** Michael C. Honigberg, MD, MPP, Massachusetts General Hospital, 185 Cambridge Street CPZN 3.190, Boston, MA 02114.

## Abstract

Clonal hematopoiesis of indeterminate potential (CHIP) represents the age-related expansion of hematopoietic stem cells with preleukemic mutations. However, its association with all-cause and cause-specific mortality has not been well characterized in older adults. We aimed to evaluate whether CHIP is associated with all-cause and cause-specific mortality in a population of older women in the United States. Our study included 6,704 participants in the Women’s Health Initiative Long Life Study (WHI-LLS) without hematologic malignancy. The co-primary exposures were any CHIP (variant allele frequency [VAF] ≥ 2%) and large CHIP (VAF ≥ 10%), and the primary outcome was all-cause mortality. Multivariable-adjusted Cox proportional hazards models tested the associations of CHIP and CHIP subtypes with all-cause and cause-specific mortality. Any CHIP and large CHIP were independently associated with all-cause mortality, with multivariable-adjusted hazard ratios (aHRs) of 1.12 (95% confidence interval [CI] 1.04-1.21; *P* = 0.003) and 1.28 (95% CI 1.15-1.43; *P* < 0.001), respectively. In gene-specific analyses, non-*DNMT3A* CHIP was associated with all-cause mortality (aHR: 1.22 [95% CI: 1.12-1.34], *P* < 0.001), while *DNMT3A* CHIP was not (aHR: 1.07 [95% CI: 0.98-1.18], *P* = 0.13). Furthermore, large CHIP was associated with cardiovascular (aHR: 1.29 [95% CI: 1.08-1.55], *P* = 0.006), cancer (aHR: 1.49 [95% CI: 1.11-2.02], *P* = 0.009), and neurologic (aHR: 1.40 [95% CI: 1.07-1.84], *P* = 0.02) death. In this cohort of older women, CHIP, particularly large clones and non-*DNMT3A* CHIP, was associated with all-cause and cause-specific mortality. These findings suggest that clonal size and subtype may differentially influence mortality risk.

## Introduction

Clonal hematopoiesis of indeterminate potential (CHIP) is characterized by the acquisition of somatic mutations that drive clonal expansion of hematopoietic stem cells, in the absence of cytopenias or dysplastic hematopoiesis.^1^ CHIP most commonly arises from mutations in genes involved in epigenetic and transcriptional regulation (e.g., *DNMT3A, TET2, ASXL1, JAK2*) as well as DNA damage repair (e.g., *PPM1D, TP53*) and spliceosome genes (e.g., *SF3B1, SRSF2*). Conventionally, CHIP is defined by the presence of a somatic variant with a variant allele frequency (VAF) of 2% or higher in peripheral blood. The prevalence of CHIP increases with age, affecting approximately 10-20% of individuals older than 70 years.^2^

The presence of CHIP has been associated with a 40% increase in all-cause mortality compared with individuals without CHIP in midlife cohorts.^3^ CHIP is also associated with an increased risk of cardiovascular disease (CVD)—including arrhythmias^4^, heart failure^5^, and atherosclerotic CVD^6,7^—with consistently stronger associations reported for *TET2* CHIP carriers in particular.^8^ By contrast, evidence suggests that CHIP may be protective against Alzheimer’s disease, a condition that predominantly affects older individuals.^9^ Although some studies have distinguished between cancer-related and non-cancer-related deaths due to CHIP, there is a lack of studies examining other specific causes of mortality.^10^ Overall, the associations between CHIP and its gene-specific subtypes with all-cause and other causes of mortality remain incompletely characterized, especially in older populations where CHIP prevalence and mortality risk are both highest. In this study, we evaluated whether CHIP and gene-specific CHIP subtypes were associated with all-cause and specific causes of death in older adults in the Women’s Health Initiative (WHI).

## Methods

### Study Cohort

The WHI is a long-term, prospective study of 161,808 postmenopausal women aged 50 to 79 years recruited between 1993 and 1998 across 40 geographically diverse clinical centers in the United States (US). At enrollment, participants completed self-administered questionnaires covering demographics; general health; clinical characteristics; functional status; lifestyle behaviors; reproductive, medical, and family history; personal habits; medication use; and dietary intake. Detailed descriptions of the study design, eligibility criteria, recruitment methods, and measurement protocols have been described elsewhere.^11^

The WHI Long Life Study (LLS) consisted of a one-time in-person home visit focused on participants between ages 63 and 98, conducted between March 2012 and May 2013. This visit, occurring 14 to 19 years after initial enrollment, included blood sample collection and assessments of clinical and functional status. It included former participants of the hormone therapy trial and other Black and Hispanic/Latina participants, excluding those residing in an institution or unable to provide informed consent. Of 7,875 participants who completed the LLS visit, 7,481 underwent a successful blood draw, and 6,976 individuals underwent CHIP targeted sequencing.^12^ After further exclusions for prevalent hematologic malignancies, lack of follow-up data after the LLS visit, and missing key covariate data, 6,704 participants were retained in the present study (**Figure 1**).

**Figure 1.**
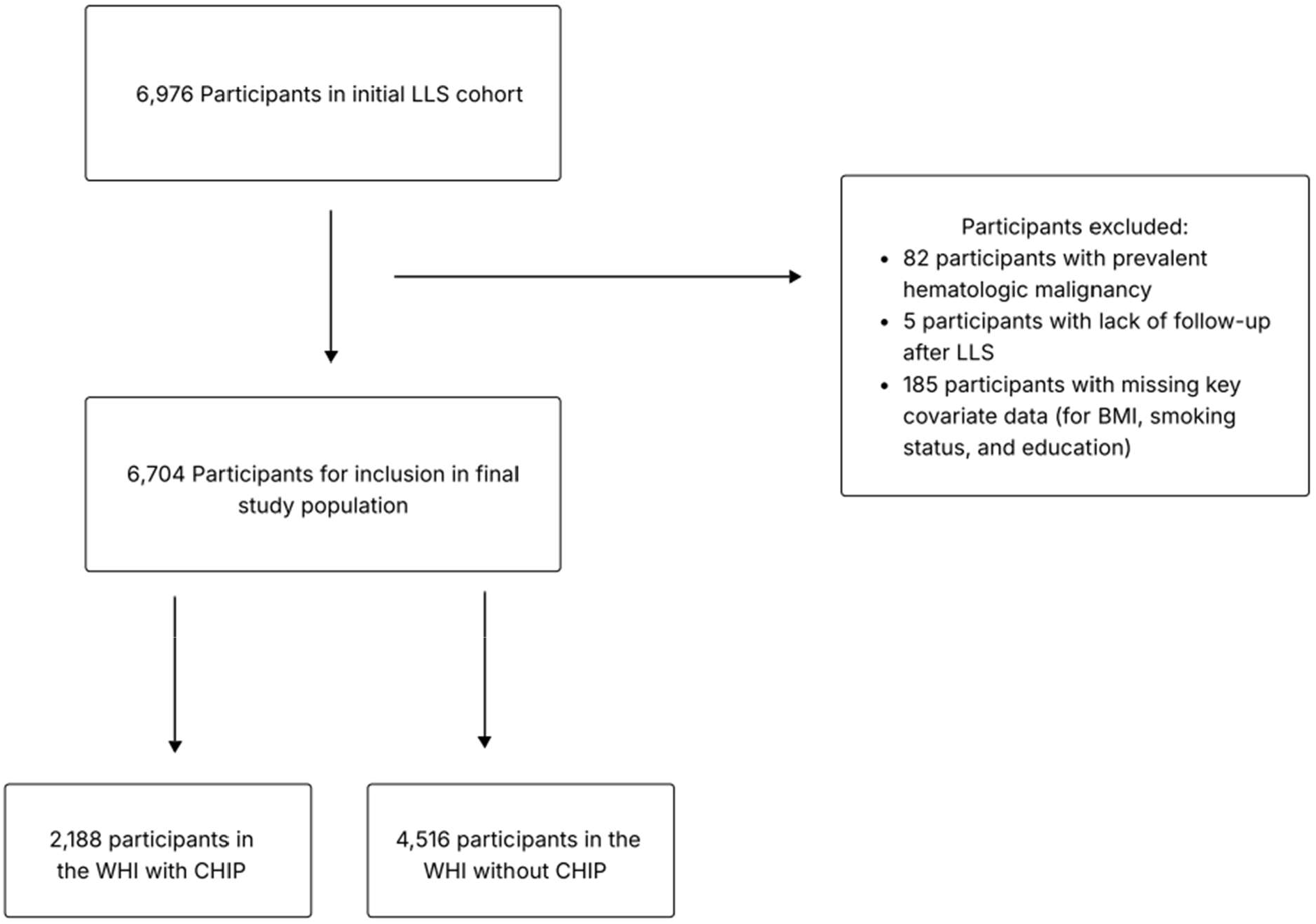
Study Flow Diagram. The study cohort included participants from the Women’s Health Initiative (WHI) Long Life Study (LLS) with genomic DNA extracted at the LLS visit. Participants were excluded for missingness of key covariates (body mass index [BMI], smoking status, and education), prevalent hematologic malignancies, or lack of follow-up after the LLS visit.

### CHIP detection

CHIP mutations were identified using single-molecule molecular inversion probe sequencing (smMIPS).^13^ The smMIPS capture panel targeted coding exons (±5 base pairs) of 11 common CHIP genes (*ASXL1, CBL, DNMT3A, GNB1, PPM1D, SF3B1, TET2, TP53, U2AF1, ZBTB33*, and *ZNF318*), along with recurrent mutational hotspots in 4 additional genes (*SRSF2, IDH1, IDH2*, and *JAK2*), as described in detail previously.^14^ The median depth of coverage in the present study cohort was 4,586x (interquartile range [IQR], 3,250-6,096x).

### Exposures and outcomes

The co-primary study exposures were the presence of any CHIP at the LLS visit, defined as CHIP with VAF ≥ 2%, and large CHIP, defined as CHIP with VAF ≥ 10%. *DNMT3A*, non-*DNMT3A, TET2, ASXL1, JAK2*, spliceosome (*SRSF2* and *SF3B1*), and DNA damage repair (DDR; *PPM1D* and *TP53*) CHIP were included as secondary exposures. For all analyses, the reference group consisted of participants without any CHIP (VAF < 2%).

The at-home LLS visit was conducted by a paramedical professional and included anthropometric measurements, blood pressure, medical history, medications, and phlebotomy for CVD biomarkers. Race, ethnicity, education, and smoking status were categorized based on self-reported identification. Education was categorized as either attainment of any higher education or, at most, a high school diploma or General Educational Development (GED) credential. Smoking status was categorized as “ever” or “never” smoking. Participants with missing covariate values (<2% missingness) were excluded.

Outcomes were ascertained using standardized physician review, classification, and adjudication based on underlying cause of death codes. The primary outcome was all-cause mortality. Secondary outcomes were the most common types of known non-accidental cause-specific mortality: cardiovascular, cancer-related, neurologic, and infectious mortality (**Supplemental Table 1**).

### Statistical analysis

Participant characteristics were compared between those with and without CHIP. Continuous participant characteristics were compared using t-tests for normally distributed variables and using Wilcoxon rank-sum tests for non-normally distributed variables. Categorical participant characteristics were compared using the Pearson chi-squared test.

In primary analyses, Cox proportional hazards models tested the associations of any CHIP and large CHIP with all-cause mortality. Minimally adjusted models (Model 1) accounted for age, race, and ethnicity. Fully adjusted models (Model 2) additionally incorporated education, smoking status, and body mass index (BMI). Secondary analyses of gene-specific CHIP subtypes and cause-specific mortality used the same fully adjusted models.

Several sensitivity analyses were conducted to evaluate the robustness of our findings. First, Fine-Gray competing risks regression accounted for competing causes of death for cause-specific mortality. Second, analyses were repeated after excluding participants with prevalent CVD or cancer. Third, inverse probability selection weights were applied to account for potential survivor bias related to participation in the LLS cohort. Finally, additional models evaluated whether the associations between CHIP and cardiovascular mortality were independent of traditional cardiovascular risk factors.

Two-sided *P* < 0.05 was considered statistically significant for the primary analyses. Findings from secondary analyses should be considered supportive and hypothesis-generating, given the possibility of type I error. All analyses were performed in R version 4.3.1.

## Results

### Participant Characteristics

The final analytic cohort included 6,704 participants (**Figure 1**), of whom 2,188 (32.6%) had CHIP at the LLS visit (**Table 1**). The most commonly mutated genes included *DNMT3A* (n = 1,269 [18.9%]), followed by *TET2* (n = 654 [9.8%]), DDR (n = 202 [3.0%]), spliceosome (n = 97 [1.4%]), *ASXL1* (n = 91 [1.4%]), and *JAK2* (n = 56 [0.8%]).

The mean (standard deviation [SD]) age of participants was 79.3 (6.8) years, with 4,217 (62.9%) self-identifying as White. Participants with CHIP were more likely to be older and self-identify as White (**Table 1**). CHIP carriers also had higher systolic blood pressure as well as lower total and LDL cholesterol relative to non-CHIP carriers.

**Table 1.** Participant Characteristics in the Women’s Health Initiative (WHI) Long Life Study (LLS).

| Variable | Overall (n = 6,704) | No CHIP (n = 4,516) | Any CHIP (n = 2,188) | P-value |
| --- | --- | --- | --- | --- |
| Age at LLS | 79.3 ± 6.8 | 78.6 ± 6.8 | 80.9 ± 6.5 | <0.001 |
| Self-reported race |  |  |  |  |
| White | 4217 (62.9%) | 2754 (61.0%) | 1463 (66.9%) | <0.001 |
| non-White | 2487 (37.1%) | 1762 (39.0%) | 725 (33.1%) |  |
| Self-reported ethnicity (Hispanic/Latina) | 1134 (16.9%) | 826 (18.3%) | 308 (14.1%) | <0.001 |
| Education ≥ Higher education | 5242 (78.2%) | 3543 (78.5%) | 1699 (77.7%) | 0.47 |
| Smoking status (ever vs. never) | 3018 (45.0%) | 2004 (44.4%) | 1014 (46.3%) | 0.14 |
| Body mass index, kg/m<sup>2</sup> | 28.9 ± 5.9 | 29.0 ± 6.1 | 28.7 ± 5.5 | 0.06 |
| Systolic blood pressure, mm Hg | 124.0 (117.0-134.0) | 124.0 (117.0-134.0) | 125.0 (118.0-134.5) | 0.01 |
| Diastolic blood pressure, mm Hg | 72.0 (67.0-79.0) | 72.0 (67.0-79.0) | 72.0 (66.0-79.0) | 0.53 |
| Total cholesterol, mg/dL | 194.0 (169.0-222.0) | 196.0 (170.0-223.0) | 191.0 (168.2-220.0) | 0.01 |
| HDL cholesterol, mg/dL | 59.0 (50.0-69.0) | 59.0 (50.0-69.0) | 58.0 (50.0-69.0) | 0.10 |
| LDL cholesterol, mg/dL | 111.0 (90.0-136.0) | 112.0 (90.0-137.0) | 109.0 (90.0-135.0) | 0.02 |
| Cholesterol-lowering medication use | 2745 (40.9%) | 1889 (41.8%) | 856 (39.1%) | 0.04 |
| Hypertension | 5327 (79.5%) | 3613 (80.0%) | 1714 (78.3%) | 0.12 |
| Anti-hypertensive medication use | 4008 (59.8%) | 2674 (59.2%) | 1334 (61.0%) | 0.18 |
| Diabetes mellitus | 2165 (32.3%) | 1445 (32.0%) | 720 (32.9%) | 0.47 |
| C-reactive protein, mg/dL | 1.8 (0.9-3.8) | 1.8 (0.9-3.8) | 1.8 (0.9-3.7) | 0.92 |

Values are mean ± standard deviation (SD), median (interquartile range [IQR]), or n (%). Participant characteristics were compared between individuals with and without clonal hematopoiesis of indeterminate potential (CHIP). Continuous variables were compared using the Student’s t-test for normally distributed variables and the Wilcoxon rank-sum test for non-normally distributed variables. Categorical variables were compared using the Pearson chi-square test.

### Mortality Rates by CHIP Status

Over a median (IQR) 10.1 (7.0-10.8) years of follow-up, 2,911 (43.4%) individuals died. Participants without CHIP had 44.79 deaths per 1,000 person-years (95% CI, 42.77-46.91), those with any CHIP had 61.74 deaths per 1,000 person-years (95% CI, 58.22-65.47), and those with large CHIP had 73.97 deaths per 1,000 person-years (95% CI, 66.93-81.76) (**Table 2**). Among CHIP carriers, participants with *DNMT3A* CHIP had the lowest mortality rate of 55.86 deaths per 1,000 person-years (95% CI, 51.57-60.51), while participants with *JAK2* CHIP had the highest mortality rate at 104.72 deaths per 1,000 person-years (95% CI, 76.81-142.76).

**Table 2.** Associations between CHIP and All-Cause Mortality in the WHI LLS.

| CHIP Subtype | N (%) with subtype | Deaths, n (%) | Mortality Rate, Deaths per 1,000 Person-years (95% CI) | Model 1 aHR (95% CI) | P-value | Model 2 aHR (95% CI) | P-value |
| --- | --- | --- | --- | --- | --- | --- | --- |
| No CHIP | 4516 (67.4%) | 1796 (39.8%) | 44.79 (42.77-46.91) | Ref. | Ref. | Ref. | Ref. |
| Any CHIP | 2188 (32.6%) | 1115 (51.0%) | 61.74 (58.22-65.47) | 1.13 (1.05-1.22) | 0.002 | 1.12 (1.04-1.21) | 0.003 |
| Large CHIP | 664 (9.9%) | 384 (57.8%) | 73.97 (66.93-81.76) | 1.29 (1.16-1.44) | <0.001 | 1.28 (1.15-1.43) | <0.001 |
| *DNMT3A* CHIP | 1269 (18.9%) | 602 (47.4%) | 55.86 (51.57-60.51) | 1.08 (0.99-1.19) | 0.09 | 1.07 (0.98-1.18) | 0.13 |
| Non-*DNMT3A* CHIP | 1183 (17.6%) | 675 (57.1%) | 72.31 (67.06-77.98) | 1.23 (1.12-1.34) | <0.001 | 1.22 (1.12-1.34) | <0.001 |
| *TET2* CHIP | 654 (9.8%) | 366 (56.0%) | 70.08 (63.26-77.64) | 1.19 (1.06-1.33) | 0.003 | 1.19 (1.06-1.33) | 0.003 |
| *ASXL1* CHIP | 91 (1.4%) | 53 (58.2%) | 75.27 (57.50-98.52) | 1.21 (0.92-1.59) | 0.18 | 1.22 (0.93-1.60) | 0.16 |
| *JAK2* CHIP | 56 (0.8%) | 40 (71.4%) | 104.72 (76.81-142.76) | 2.13 (1.55-2.91) | <0.001 | 2.04 (1.49-2.79) | <0.001 |
| Spliceosome CHIP | 97 (1.4%) | 62 (63.9%) | 80.35 (62.65-103.06) | 1.26 (0.97-1.62) | 0.08 | 1.24 (0.96-1.60) | 0.10 |
| DDR CHIP | 202 (3.0%) | 119 (58.9%) | 75.50 (63.08-90.36) | 1.26 (1.05-1.52) | 0.01 | 1.25 (1.04-1.50) | 0.02 |

Associations between clonal hematopoiesis of indeterminate potential (CHIP) subtypes and all-cause mortality were assessed using Cox proportional hazards models. Model 1 was adjusted for age, race, and ethnicity. Model 2 was adjusted for age, race, ethnicity, education, smoking status, and BMI. The reference group consisted of individuals with no CHIP. Large CHIP was defined as variant allele frequency (VAF) $\geq 10\%$ . aHR = adjusted hazard ratio; CI = confidence interval; WHI = Women's Health Initiative; LLS = Long Life Study.

Cardiovascular mortality was the most frequent cause of death for CHIP carriers and non-CHIP carriers based on both mortality rates and cumulative incidence (**Supplemental Tables 2** and **3**). Individuals generally demonstrated the highest absolute mortality rate differences for cardiovascular mortality, except for DDR CHIP, for which the highest absolute mortality rate difference was due to infectious causes (**Supplemental Table 4**).

### Associations of CHIP with All-Cause Mortality

In models adjusted for age, race, and ethnicity, any CHIP and large CHIP were associated with all-cause mortality, with aHRs of 1.13 (95% CI, 1.05-1.22; *P* = 0.002) and 1.29 (95% CI, 1.16-1.44; *P* < 0.001), respectively (**Table 2**). After further adjustment for education, smoking status, and BMI, associations with all-cause mortality were materially unchanged; associations were similarly observed for any CHIP (aHR: 1.12 [95% CI: 1.04-1.21]; *P* = 0.003) and large CHIP (aHR: 1.28 [95% CI: 1.15-1.43]; *P* < 0.001).

In fully adjusted models, non-*DNMT3A* CHIP was associated with all-cause mortality (aHR: 1.22 [95% CI: 1.12-1.34], *P* < 0.001), driven by associations with *TET2* (aHR: 1.19 [95% CI: 1.06-1.33], *P* = 0.003), *JAK2* (aHR: 2.04 [95% CI: 1.49-2.79], *P* < 0.001), and DDR CHIP (aHR: 1.25 [95% CI: 1.04-1.50], *P* = 0.02). In contrast, *DNMT3A* CHIP was not associated with all-cause mortality (aHR: 1.07 [95% CI: 0.98-1.18], *P* = 0.13; **Table 2**).

Results were consistent across sensitivity analyses that (1) incorporated inverse probability selection weights to account for potential survivor bias in the LLS cohort, and (2) excluded individuals with prevalent CVD or cancer to evaluate whether the association between CHIP and mortality persisted in a lower-disease burden population (**Supplemental Tables 5** and **6**).

### Associations of CHIP with Cause-Specific Mortality

In fully adjusted models, the presence of large CHIP was associated with an increased risk of cardiovascular mortality, with an aHR of 1.28 (95% CI, 1.06-1.53; *P* = 0.008), while any CHIP was associated with a borderline increase in risk (aHR, 1.12 [95% CI, 0.99-1.27]; *P* = 0.08) (**Table 3**). Non-*DNMT3A, JAK2*, and spliceosome CHIP were also associated with increased cardiovascular mortality, with aHRs of 1.24 (95% CI, 1.07-1.43; *P* = 0.004), 2.77 (95% CI, 1.77-4.34; *P* < 0.001), and 1.53 (95% CI, 1.05-2.22; *P* = 0.03), respectively.

**Table 3.** Associations between CHIP and Cause-Specific Mortality in WHI LLS.

| CHIP Subtype | Cardiovascular |  |  | Cancer |  |  | Neurologic |  |  | Infectious |  |  |
| --- | --- | --- | --- | --- | --- | --- | --- | --- | --- | --- | --- | --- |
|  | Events, n (%) | aHR (95% CI) | P-value | Events, n (%) | aHR (95% CI) | P-value | Events, n (%) | aHR (95% CI) | P-value | Events, n (%) | aHR (95% CI) | P-value |
| No CHIP | 638 (14.1%) | Ref. | Ref. | 242 (5.4%) | Ref. | Ref. | 267 (5.9%) | Ref. | Ref. | 131 (2.9%) | Ref. | Ref. |
| Any CHIP | 413 (18.9%) | 1.12 (0.99-1.27) | 0.08 | 156 (7.1%) | 1.29 (1.05-1.58) | 0.02 | 174 (8.0%) | 1.13 (0.94-1.37) | 0.20 | 73 (3.3%) | 1.00 (0.75-1.33) | 1.00 |
| Large CHIP | 145 (21.8%) | 1.28 (1.06-1.53) | 0.008 | 54 (8.1%) | 1.52 (1.13-2.05) | 0.006 | 65 (9.8%) | 1.40 (1.06-1.83) | 0.02 | 18 (2.7%) | 0.82 (0.50-1.34) | 0.42 |
| *DNMT3A* CHIP | 217 (17.1%) | 1.06 (0.91-1.24) | 0.45 | 95 (7.5%) | 1.32 (1.04-1.67) | 0.02 | 100 (7.9%) | 1.20 (0.95-1.51) | 0.12 | 33 (2.6%) | 0.79 (0.54-1.16) | 0.23 |
| Non-*DNMT3A* CHIP | 260 (22.0%) | 1.24 (1.07-1.43) | 0.004 | 89 (7.5%) | 1.39 (1.08-1.78) | 0.01 | 94 (7.9%) | 1.07 (0.84-1.35) | 0.59 | 46 (3.9%) | 1.16 (0.82-1.63) | 0.41 |
| *TET2* CHIP | 141 (21.6%) | 1.20 (1.00-1.45) | 0.05 | 50 (7.6%) | 1.40 (1.03-1.91) | 0.03 | 53 (8.1%) | 1.08 (0.80-1.45) | 0.61 | 24 (3.7%) | 1.08 (0.70-1.68) | 0.73 |
| *ASXL1* CHIP | 22 (24.2%) | 1.33 (0.87-2.04) | 0.19 | 8 (8.8%) | 1.59 (0.79-3.23) | 0.2 | 7 (7.7%) | 1.00 (0.47-2.12) | 1.00 | 4 (4.4%) | 1.28 (0.47-3.47) | 0.63 |
| *JAK2* CHIP | 20 (35.7%) | 2.77 (1.77-4.34) | <0.001 | 4 (7.1%) | 1.47 (0.55-3.96) | 0.45 | 6 (10.7%) | 2.13 (0.95-4.81) | 0.07 | 1 (1.8%) | 0.71 (0.10-5.08) | 0.73 |
| Spliceosome CHIP | 29 (29.9%) | 1.53 (1.05-2.22) | 0.03 | 4 (4.1%) | 0.70 (0.26-1.88) | 0.47 | 11 (11.3%) | 1.43 (0.78-2.61) | 0.25 | 3 (3.1%) | 0.82 (0.26-2.57) | 0.73 |
| DDR CHIP | 30 (14.9%) | 0.84 (0.58-1.21) | 0.34 | 17 (8.4%) | 1.56 (0.95-2.56) | 0.08 | 15 (7.4%) | 1.00 (0.60-1.69) | 0.99 | 16 (7.9%) | 2.35 (1.39-3.97) | 0.001 |

Associations between clonal hematopoiesis of indeterminate potential (CHIP) subtypes and cause-specific mortality were assessed using Cox proportional hazards models. All models were adjusted for age, race, ethnicity, education, smoking status, and BMI. Large CHIP was defined as a variant allele frequency (VAF) $\geq 10\%$ . aHR = adjusted hazard ratio; CI = confidence interval; WHI = Women's Health Initiative; LLS = Long Life Study. Cells with “–” indicate that no one with that CHIP subtype died from that specific cause. Causes of death were classified as cardiovascular, cancer, neurologic, and infectious mortality.

The presence of any CHIP and large CHIP was associated with increased cancer mortality, with aHRs of 1.29 (95% CI, 1.05-1.58; *P* = 0.02) and 1.52 (95% CI, 1.13-2.05; *P* = 0.006), respectively (**Table 3**). Participants with *DNMT3A* and *TET2* CHIP also had an increased risk of cancer mortality, with aHRs of 1.32 (95% CI, 1.04-1.67; *P* = 0.02) and 1.40 (95% CI, 1.03-1.91; *P* = 0.03), respectively.

The presence of large CHIP was associated with increased risk of neurologic mortality, with an aHR of 1.40 (95% CI, 1.06-1.83; *P* = 0.02), while any CHIP was not (**Table 3**). In analyses of specific causes of neurologic death, large CHIP and *JAK2* CHIP were associated with increased Alzheimer’s disease-related mortality (**Supplemental Table 7**), while no associations were observed for dementia not specified as Alzheimer’s-related or Parkinson’s disease.

The presence of any CHIP or large CHIP was not associated with infectious mortality (**Table 3**). However, individuals with DDR CHIP had an over two-fold increased risk of infectious mortality (aHR: 2.35 [95% CI: 1.39-3.97]; *P* = 0.001).

Results were broadly consistent across sensitivity analyses that (1) incorporated inverse probability selection weights, (2) excluded individuals with prevalent CVD or cancer, and (3) used competing risk regression models (**Supplemental Tables 8-10**). However, after excluding participants with prevalent CVD or cancer, no associations were observed between any CHIP subgroup and cancer-related mortality (**Supplemental Table 9**). Finally, in a sensitivity analysis additionally adjusting for cardiovascular risk factors (systolic blood pressure, LDL cholesterol, diabetes treatment, and C-reactive protein), associations between any CHIP and large CHIP with cardiovascular mortality no longer remained statistically significant, while non-*DNMT3A, TET2*, and *ASXL1* CHIP remained associated with increased cardiovascular mortality (**Supplemental Table 11**).

## Discussion

In this prospective cohort of older women, CHIP was common and was independently associated with increased risk of all-cause mortality, with particularly strong associations observed for large CHIP and several gene-specific subtypes. These associations were largely driven by non-*DNMT3A* mutations, including *TET2, JAK2*, and DDR genes, whereas *DNMT3A* CHIP was not independently associated with mortality. In cause-specific analyses, we observed associations between large CHIP and cardiovascular, cancer, and neurologic mortality. Together, these findings highlight heterogeneity in the prognostic implications of CHIP in older adults and suggest that both clone size and mutation class may influence long-term mortality risk.

We can draw several conclusions based on these results. First, our results align with prior reported associations between CHIP and all-cause mortality and extend these findings to an older female population. Previously, Jaiswal et al. reported that CHIP was associated with a hazard ratio of 1.40 for all-cause mortality in a pooled cohort of 17,182 midlife participants (median age of 58 years).^3^ In our study, we found that any CHIP and large CHIP were associated with 12% and 28% increased risk of all-cause mortality, respectively, suggesting stronger associations with larger clonal burden. Overall, our results confirm previously observed associations between CHIP and all-cause mortality in an older population.^7,15^

Second, the association of CHIP with mortality varies depending on the specific driver mutation. We observed strong associations between non-*DNMT3A, TET2*, and *JAK2* CHIP and all-cause mortality. *TET2* CHIP warrants particular attention as the most common subtype of non-*DNMT3A* CHIP, with established relevance to atherosclerotic CVD and heart failure.^6,16^ Prior experimental work supports the premise that *TET2* mutations are causally associated with atherosclerosis.^17^ *TET2*^+/−^ macrophages have demonstrated increased LDLR expression and lipid uptake *in vitro*, supporting the role of upregulated lipid metabolism and inflammatory pathways in mortality risk.^8^ One potential mechanism is that in aging populations, elevated levels of IL-1 in bone marrow may drive *TET2*^+/−^ clonal expansion through increased proliferation of hematopoietic stem cells.^18^ *TET2* may also play a role in the development of hematologic malignancies through its role in promoting DNA instability and stem cell self-renewal.^19^ Collectively, these findings suggest that *TET2* CHIP promotes a systemic pro-inflammatory state that may contribute to multi-organ aging and increased all-cause mortality.

Cardiovascular mortality accounted for the largest share of CHIP-associated deaths, with *JAK2* mutations conferring nearly 3-fold relative risk to those without CHIP. This finding is consistent with prior studies linking *JAK2* CHIP to CVD.^20^ Constitutive JAK/STAT signaling can promote expression of inflammatory cytokines and may increase oxidative stress, pathways implicated in atherosclerosis and adverse outcomes following myocardial infarction.^21,22^ In addition, hyperactivation of *JAK2* may disrupt cholesterol efflux in macrophages and lead to accumulation of foam cells in plaques, potentially accelerating atherogenesis.^23^ *JAK2* V617F has been linked to enhanced platelet activation and crosstalk and arterial thrombosis.^24^ These findings support a model in which *JAK2* CHIP promotes cardiovascular mortality through both atherothrombotic and myocardial pathways and raise the possibility that targeted inhibition of the *JAK2* pathway could mitigate cardiovascular risk in individuals with CHIP, a hypothesis that warrants prospective evaluation.

Furthermore, we identified a significant association between large CHIP clones and neurologic mortality. This finding contrasts with previous work reporting a protective effect of CHIP for individuals with dementia and an association between larger CHIP clones and reduced dementia in postmenopausal women.^9,25^ One potential explanation for these seemingly discordant findings is that CHIP may differentially influence the development of neurologic disease versus vulnerability to death once disease or age-related decline is present. Upregulated inflammatory gene expression driven by CHIP mutations may promote a chronic, low-grade systemic inflammation, known as “inflammaging,” which has been implicated in age-related conditions such as sarcopenia, osteoporosis, and frailty,^26,27^ thereby contributing to mortality risk in older adults. Recent work further supports a direct role for clonal somatic mutations in neurodegeneration, demonstrating that *TET2, ASXL1*, and *DNMT3A* variants are enriched in microglia-like macrophages in Alzheimer’s disease and potentially drive neuroinflammation.^28,29^ Taken together, these findings raise the possibility that large CHIP clones may increase neurologic mortality both by contributing to neuroinflammatory disease processes and by reducing physiologic resilience in older adults, although further studies are needed to clarify these mechanisms.

Finally, we found an association between DDR CHIP and infectious mortality, which remained consistent after excluding patients with prevalent cancer, suggesting that it is unlikely to be explained solely by comorbidity confounding. Dysregulated immune responses arising from these mutant clones could plausibly impair host defense or promote maladaptive inflammation during infection, thereby increasing infection-related mortality. However, the biological basis of this association remains unclear and warrants further investigation in experimental and prospective cohort studies.

Strengths of our study include the use of high-coverage targeted sequencing, which enabled the detection of CHIP with sensitivity for low-VAF clones that would have otherwise been missed by conventional whole genome or whole exome sequencing approaches, and a focus on older individuals, who have the highest absolute risk for mortality.^14^ In addition, the relatively large sample size and extended follow-up of the WHI-LLS provided statistical power to examine associations with CHIP subtypes. However, this study also has several limitations. First, our sample size was modest for gene-specific analyses beyond the most common mutations (*DNMT3A* and *TET2*), potentially limiting our ability to detect associations with rarer CHIP subtypes. Our analyses were also limited by relatively small numbers of events for neurologic and infectious mortality for some CHIP subtypes. The study also consisted exclusively of older women selected for longevity, which may limit generalizability to men and younger adults.

Relatedly, our findings may be impacted by survivor bias, as individuals who developed CHIP earlier in life and experienced adverse outcomes may have been less likely to survive to LLS enrollment. However, results were consistent in sensitivity analyses incorporating inverse probability selection weights. Finally, we did not adjust for multiple comparisons for the primary analyses in our study as we pre-specified two highly correlated co-primary exposures, any CHIP and large CHIP, and a primary outcome, all-cause mortality.

## Conclusions

In this population of older women, CHIP was associated with increased risk of all-cause mortality, particularly among individuals with non-*DNMT3A* mutations and larger clones. Large CHIP was associated with increased cardiovascular, cancer, and neurologic mortality. These findings underscore the heterogeneity of risk across CHIP subtypes and suggest that both mutation class and clonal size may help refine prognostic assessment in older adults. Future studies are needed to elucidate the biological mechanisms linking CHIP to adverse outcomes and to determine whether targeted interventions could mitigate CHIP-associated morbidity and mortality.

## Acknowledgements

The authors would like to thank the participants of the Women’s Health Initiative Long Life Study.

## Authorship Contributions

A.C., D.E., and M.C.H. conceived and designed the study. A.C. and D.E. conducted formal analysis and drafted the original manuscript. All authors (A.C., D.E., M.M.U., Y.P. J.C., J.O.K., S.J., P.D., A.H.S., G.L.A., R.C., R.W., J.W., A.G.B., P.N., C.K., M.M., E.A.W., J.E.M., A.P.R., and M.C.H.) contributed to critical revision of the manuscript. M.C.H. provided direct supervision and project administration. The Women’s Health Initiative (WHI) investigators (A.G.B., C.K., M.J.M., E.A.W., J.E.M., A.P.R., J.E.M., A.P.R., and M.C.H.) contributed to overall supervision of the research. All authors reviewed and approved the final version of the manuscript and agree to be accountable for the work.

## Data sharing statement

WHI data and documentation are available from the WHI Clinical Coordinating Center (https://www.whi.org/get-started or).

## Code availability

Scripts for CHIP calling and filtering are available at https://github.com/MMesbahU/clonalhematopoiesis-in-whi.

## Clinical trial number

ClinicalTrials.gov ID NCT00000611.

## Ethical approval

The WHI project was reviewed and approved by the Fred Hutchinson Cancer Research Center Institutional Review Board in accordance with the U.S. Department of Health and Human Services regulations at 45 CFR 46 (approval number: IR# 3467-EXT). Participants provided written informed consent to participate. Additional consent to review medical records was obtained through signed written consent. The Fred Hutchinson Cancer Research Center has an approved Federal-wide Assurance on file with the Office for Human Research Protections under assurance number 0001920.

